# Effort-reward imbalance at work, glycated hemoglobin and prediabetes prevalence in a prospective cohort

**DOI:** 10.1101/2023.01.07.23284201

**Authors:** C Riopel, X Trudel, A Milot, D Laurin, M Gilbert-Ouimet, C Brisson

## Abstract

**Contex:** Prospective studies and meta-analyses suggest that psychosocial stressors at work from the effort-reward imbalance model are associated with an increased risk of type 2 diabetes mellitus (T2DM). Prediabetes is an intermediate disorder on the glucose metabolism continuum. It increases the risk of developing T2DM, while also being separately associated with increased mortality. Evidence about the effect of effort-reward imbalance at work on prediabetes is scarce.

**Objective:** The objective was to evaluate, in women and men, the association between effort-reward imbalance at work, glycated hemoglobin (HbA1c) concentration and the prevalence of prediabetes in a prospective cohort study.

**Methods:** This study was conducted among 1,354 white-collar workers followed for an average of 16 years. Effort-reward imbalance at work (ERI) was measured at baseline (1999-2001) using a validated instrument. HbA1c was assessed at follow-up (2015-18). Several covariates were considered including sociodemographics, anthropometric, and lifestyle risk factors. Differences in mean HbA1c concentration were estimated with linear models. Prediabetes prevalence ratios (PRs) were computed using Poisson regressions models.

**Results:** In women, those exposed to effort-reward imbalance at work had a higher prevalence of prediabetes (adjusted PR=1.52, 95% confidence interval: 1.01-2.29). There was no difference in HbA1c concentration among those exposed and those unexposed to an effort-reward imbalance at work.

**Conclusion:** Among women, effort-reward imbalance at work was associated with the prevalence of prediabetes. Preventive workplace interventions aiming to reduce the prevalence of effort-reward imbalance at work may be effective to reduce the prevalence of prediabetes among women.

## Introduction

Type II diabetes mellitus (T2DM) affects over 400 million individuals across the globe and is on the rise (1). T2DM increases the risk of renal, cardiovascular and neurocognitive pathologies, making it a leading cause of mortality worldwide (1). Prediabetes refers to high blood glucose that is not within diabetes range. Prediabetes is associated with a significant increase in morbidity and mortality whether individuals develop T2DM or not (2-5). A prospective study also showed a linear relationship between glycated hemoglobin (HbA1c), an indicator of glucose metabolism, and health complications including cardiovascular diseases and all-cause mortality among nondiabetic adults (6). Primary prevention strategies should therefore target modifiable risk factors associated with glucose metabolism imbalance and prediabetes, in order to delay or prevent progression to T2DM as well as adverse health outcomes associated with prediabetes itself.

Previous evidence suggests that psychosocial stressors at work are associated with an increased risk T2DM (7, 8). Two well defined and empirically supported models have been used to assess the adverse effect of psychosocial stressors at work. The demand-control (i.e. job strain) model states that stress-related ill health risks increase from the combined effects of high psychological demand and low job control (9, 10). The effort-reward imbalance model (ERI) posits that failed reciprocity between efforts invested at work and rewards obtained in return (monetary, social and organizational) induces a high-stress situation (11). Little is known about the effect of psychosocial stressors at work on prediabetes. To our knowledge, only one prospective study, conducted among 7,503 workers in Brazil has examined this effect (12) showing sex-specific associations. Indeed, this previous study has reported that, among women, job strain is associated with a 32% increase in prediabetes incidence. However, exposure to an ERI at work showed no independent association with prediabetes. This result could be explained by the relatively low participation at baseline (30%) (13) which could have led to selection bias and effect underestimation (14, 15). Moreover, the 4-year follow-up might have been too short to capture the adverse effect of ERI exposure on prediabetes. Further studies are therefore required to determine whether ERI at work is associated with the prevalence of prediabetes. The objective of the present study was to examine the association between effort-reward imbalance at work, glycated hemoglobin (HbA1c) concentration and the prevalence of prediabetes in a prospective cohort study of women and men followed for 16 years.

## Methods

### Study Design and Population

The study sample was derived from the PROspective Quebec (PROQ) cohort, which has been previously described (16). Briefly, PROQ is a prospective white-collar workers cohort initiated in 1991–1993 (T1). The 9,188 workers (48.5% women) were recruited from 19 public organizations in Quebec City, Canada. They were followed up 8 and 24 years later (T2 and T3), with good participation rates: 75% at baseline, 90% at the 8-year follow-up and 81% at the 24-year follow-up. This study was approved by the CHU de Quebec’s Research Center ethical review board. All participants, or their respondents in case of inability, provided their informed consent.

At the 24-year follow-up (T3), a convenience sample of 2,318 participants were targeted for blood tests including HbA1c concentration. Of these, 17 died before the follow-up, 86 were lost to follow up, 344 refused to participate, 367 participated by mail only, and 41 did not get adequate blood samples. Furthermore, 77 individuals did not participate at T2 (45 refusals, 15 lost to follow-up and 17 inactive workers) and were excluded from the analysis. Finally, 32 participants had missing data on ERI exposure and were further excluded. A total of 1,354 participants (649 men and 656 women) were included in the analysis.

### Effort-reward imbalance at work

Effort-reward imbalance was measured at T2 (1999-2001) using validated scales. Reward at work was measured by nine original questions from the French version (17-19) of the effort-reward imbalance scale (three of five questions from the esteem subscale, the four-item subscale of promotion prospects and salary, and the two-item subscale of job security). The questions were answered in two steps (19). The respondents were first asked to indicate whether they agreed or disagreed that the question content described an experience typical of their work situation. If they agreed, they were then asked to indicate to what extent they felt distressed by the experience with 1 = very distressed, 2 = distressed, 3 =somewhat distressed, 4 = not at all distressed (scores were inverted for positive items). The 5-point answer mode were recoded in 3 points with a range from 9 to 27, lower values indicating lower reward. Effort was measured by nine items from the validated French version of the psychological demand scale of the Job Content Questionnaire (20, 21). The 4-point answer mode were recoded in 3 points with a range from 9 to 27, with higher values indicating higher effort. The psychometric qualities of this version have been demonstrated (22). In line with previous literature, the ERI score was calculated by dividing the effort score by the reward score and ERI was categorized in the following way: participants were considered exposed to an ERI if their effort-reward ratio was _≥_1 and unexposed if their ratio was <1.

### HbA1c and prediabetes

Non fasting blood samples were taken at T3 (2015-2018) by a trained nurse following a standardized protocol. EDTA whole-blood samples were analyzed at Biron Groupe Santé inc, a certified laboratory. HbA1c was measured using the immunochemical method on an Integra platform from Roche diagnostics (coefficient of variation of 1.6%). The *American Diabetes Association* guidelines were followed, which define prediabetes as an HbA1c between 5.7% and 6.49% (23). Workers with a HbA1c concentration under these thresholds were considered to have normal glucose metabolism. Participants with a HbA1c _≥_6.5% were considered prevalent T2DM cases.

Participants who reported a previous T2DM diagnosis were 1) excluded from the HbA1c analyses and 2) categorized as prevalent T2DM cases for the prediabetes analyses. These participants with a history of T2DM are likely to have a different HbA1c value than at time of diagnosis because of their treatment, which could diminish the strength of the association between ERI at work and HbA1c.

### Covariates

The following risk factors for prediabetes were assessed at T2 (1999-2001) and retained as covariates. Age was divided into tertiles: <40 years old, 40–49 years old and 50 years old or above. Education was defined as the highest degree obtained: less than college, college, or university. Smoking status was categorized following the WHO guidelines: nonsmokers, ex-smokers and current smokers (24). Physical activity was categorized as follows, based on participants’ answers to a validated question on the duration and frequency of their physical activity: sedentary (active once or less per week), insufficiently active (active 2–3 times per week), and active (>3 physical activity sessions per week) (25). Alcohol consumption was dichotomized based on a validated question on alcohol consumption (26): 1) nondrinkers: less than 1 alcoholic beverage per week and 2) drinkers: _≥_1 alcoholic beverage per week. Long working hours were dichotomized based on a standard full-time employment schedule in Canada: 1) <41hrs/week and 2) _≥_ 41hrs/week. Clinical characteristics (blood pressure [BP], weight, height) were measured by trained employees using validated protocols (26). BP was measured following the American Heart Association protocol (27). After 5 minutes of rest, two blood pressure measurements were taken 1–2 minutes apart and the average of the two measures was used in the analyses. BP was dichotomized according to the presence or absence of hypertension (systolic BP _≥_140 mmHg or diastolic BP _≥_90 mmHg). Body mass index (BMI) was obtained by dividing the weight in kilograms by the height in metres squared and was treated as a continuous variable. Hypercholesterolemia was self-reported.

### Statistical Analysis

First, descriptive analyses were conducted. Comparisons between men and women were examined using Student’s t-tests for continuous variables and chi-square tests for categorical variables.

Crude and adjusted HbA1c mean differences according to ERI exposure as well as its individual components (efforts and rewards) were computed using linear regression analyses. Prevalence and prevalence ratio (PR) of prediabetes with 95% confidence intervals were computed using generalized linear models with a log link and a Poisson working model (28). A robust variance was used to account for the larger variance of the Poisson distribution compared to the binomial distribution. Crude and adjusted PRs were estimated. Missing data on each variable was less than 1% except for family history of T2DM (7.6%). A dummy indicator was used to prevent the exclusion of participants with missing data on this variable. According to prior literature, the impact of psychosocial stressors at work on T2DM may be partially mediated by adoption of unhealthy lifestyle habits (29-32). Therefore, sequential adjustments were used. However, given that estimates with and without lifestyle risk factors yielded similar results, only fully adjusted models are presented.

Prior studies found important sex differences in the association between psychosocial stressors at work and glucose metabolism imbalances (12, 33). Therefore, separate analyses were performed for men and women. Statistical analyses were conducted using the statistical software package SAS, version 9.4. Two tailed statistical tests with a significance level of 0.05 were used.

## Results

Table 1 presents baseline characteristics of our study sample, by sex. Compared to men, women were younger (38.9 years old, SD=4.5 vs 40.0 years old, SD=5.2). Women were less educated; 43.2% of them held a university diploma compared to 65.2% of men. More men lived with a spouse (83.6%) than women (77.3%). Women were healthier (lower prevalence of overweight individuals, hypertension and high cholesterol) and only 61.7% of them were alcohol drinkers, compared to 76.3% of men. However, more women were current smokers (17.3%) than men (12.2%). Only 2.8% of women reported long working hours compared to 9.0% of men. Lastly, there was no difference in ERI prevalence between men and women, but more men reported high efforts at work (44.1%) more frequently when compared to women (37.4%).

**Table 1.** Characteristics of the study sample at T2 (1999-2001)

|  | **MEN** | **WOMEN** | **TOTAL** |
| --- | --- | --- | --- |
| Age (years), mean (SD) | 40.0 (5.2)* | 38.9 (4.5)* | 39.4 (4.9) |
| Education, n (%) |  |  |  |
| Less than college | 44 (6.3)* | 141 (20.5)* | 185 (13.4) |
| College | 198 (28.5)* | 249 (36.2)* | 447 (32.3) |
| University | 454 (65.2)* | 297 (43.2)* | 751 (54.3) |
| Family history of T2DM, n (%) <sup>b</sup> | 129 (20.2) | 153 (23.9) | 282 (22.1) |
| Living with a spouse, n (%) | 581 (83.6)* | 530 (77.3)* | 1111 (80.5) |
| Overweight (BMI $\geq 25.0$ kg/m<sup>2</sup>), n (%) | 405 (59)* | 215 (31.7)* | 620 (45.4) |
| Hypertension, n (%) <sup>a</sup> | 106 (15.6)* | 18 (2.7)* | 124 (9.3) |
| High cholesterol, n (%) | 189 (27.2)* | 96 (13.9)* | 285 (20.6) |
| Smoking status, n (%) |  |  |  |
| Nonsmoker | 437 (62.8)* | 352 (51.2)* | 789 (57.0) |
| Ex-smoker | 174 (25.0)* | 217 (31.5)* | 391 (28.3) |
| Current smoker | 85 (12.2)* | 119 (17.3)* | 204 (14.7) |
| Alcohol drinker, n (%) <sup>c</sup> | 530 (76.3)* | 424 (61.7)* | 954 (69.0) |
| Physical activity, n (%) <sup>d</sup> |  |  |  |
| Active | 181 (26.0) | 145 (21.1) | 326 (23.6) |
| Insufficiently active | 284 (40.9) | 288 (41.9) | 572 (41.4) |
| Sedentary | 230 (33.1) | 254 (37.0) | 484 (35.0) |
| Long working hour ($\geq 41$h/wk), n (%) | 62 (9.0)* | 19 (2.8)* | 81 (5.9) |
| Effort-reward ratio $\geq 1$, n (%) | 183 (26.8) | 167 (24.8) | 350 (25.9) |
| Efforts, n (%) |  |  |  |
| Low | 152 (22.3)* | 214 (31.9)* | 366 (27.0) |
| Medium | 229 (33.6)* | 207 (30.8)* | 436 (32.2) |
| High | 301 (44.1)* | 251 (37.4)* | 552 (40.8) |
| Rewards, n (%) |  |  |  |
| High | 211 (39.0) | 203 (30.2) | 414 (30.6) |
| Medium | 253 (37.1) | 231 (34.4) | 484 (35.8) |
| Low | 218 (32.0) | 238 (35.4) | 456 (33.7) |

<sup>a</sup>Defined as a systolic blood pressure $\geq 140$ mmHg or a diastolic blood pressure $\geq 90$ mmHg
<sup>b</sup>Measured at T3
<sup>c</sup>Drinker: $\geq 1$ drink/week
<sup>d</sup>One session is defined as being physically active for at least 20 minutes. Active: $\geq 3$ sessions/week, insufficiently active: 1–2 sessions/week, sedentary: $<1$ session/week
\*Statistically significant difference between men and women ( $p < 0.05$ )

Table 2 presents mean HbA1c concentration and mean differences according to ERI, effort and reward exposures, by sex. There was no association between ERI exposure and mean HbA1c concentration in either men or women. Also, no association was observed when efforts and reward were considered separately.

**Table 2.**
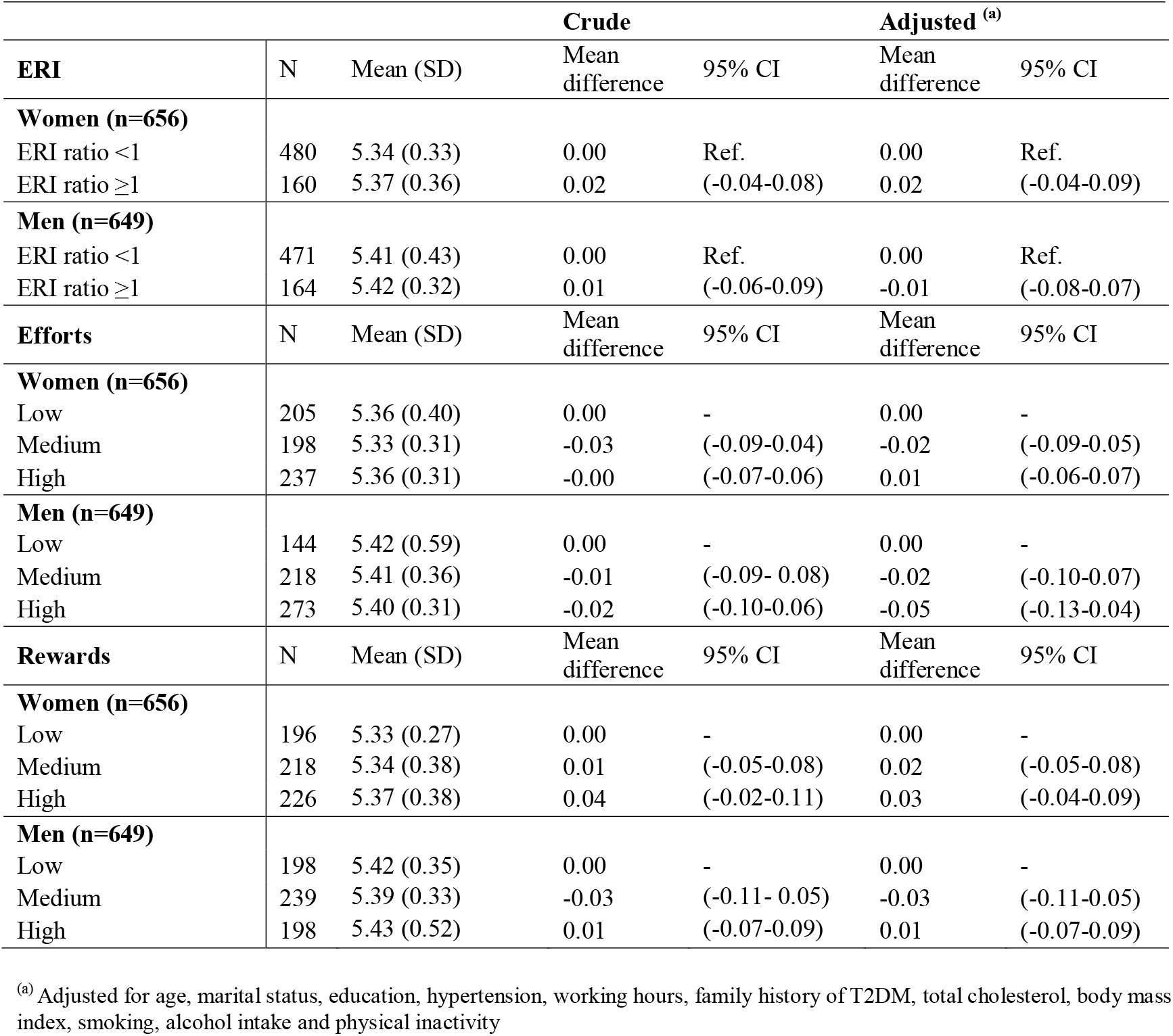
Mean HbA1c and mean difference (MD) according to ERI, effort and reward exposures, by sex

Table 3 presents the prevalence and prevalence ratio (PR) of prediabetes according to ERI, efforts and reward exposures, by sex. In women, the prevalence of prediabetes was higher among those exposed to ERI compared to unexposed women, after adjusting for potential confounders (PR=1.52, 95% CI: 1.01-2.29). In men, the results did not suggest an association between ERI exposure and prediabetes prevalence. The results did not suggest an association between efforts and prediabetes prevalence in either men or women. Lastly, in women, low rewards were associated with an increased prediabetes prevalence (PR=1.67, 95% CI: 1.02-2.72), although statistical significance was not reached in the adjusted model (PR=1.57, 95% CI: 0.94-2.60).

**Table 3.**
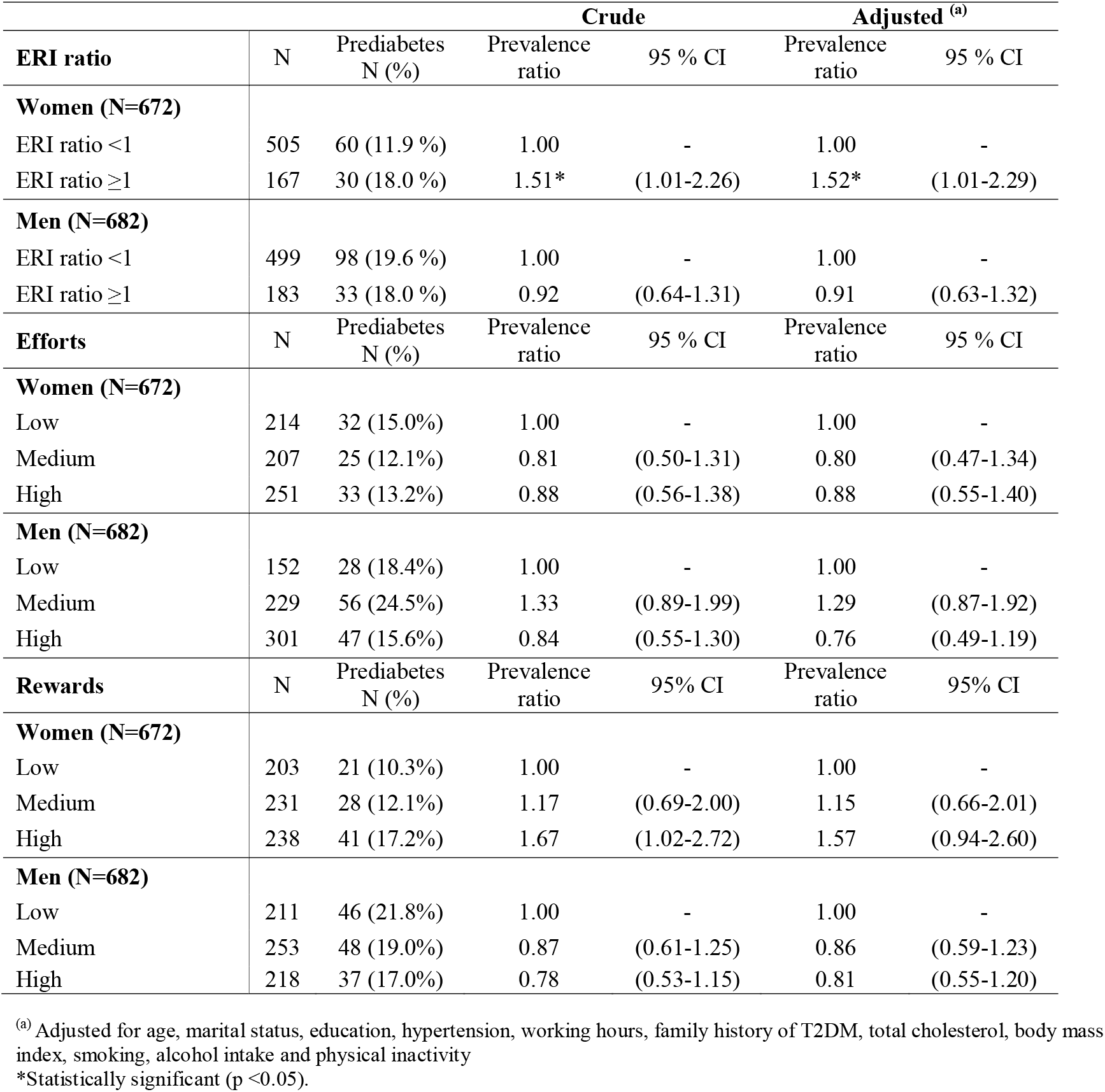
Prevalence of prediabetes and prevalence ratio (PR) according to ERI, effort and reward exposures, by sex.

## Discussion

The aim of the present study was to evaluate the association between effort-reward imbalance at work, HbA1c concentration and prediabetes prevalence in a prospective cohort study. Results showed that the prevalence of prediabetes was higher in women exposed to ERI at work, compared to unexposed women. This association was robust to adjustment for sociodemographics, anthropometrics and lifestyle risk factors. Low rewards at work, considered separately, was also associated with an increased prevalence of prediabetes among women.

Evidence about the adverse effect of psychosocial stressors at work on prediabetes is scarce. Previous cross-sectional studies on ERI and prediabetes reported conflicting results, which could be attributable to small sample sizes (34, 35). The sole prospective cohort study on ERI and prediabetes reported no association (12). The relatively low participation rate (30%) at baseline in this cohort (13) could have led to a healthy worker effect and could partly explain this null finding. Indeed, the healthy worker effect arises when less healthy workers are less likely to participate when compared to healthier workers (36). This selection bias can result in an underestimation of the true effect. Furthermore, the 4-year follow-up might not have been the optimal temporal window to capture the adverse effect of ERI on prediabetes. Indeed, previous studies have reported that an induction period of several years can be required for psychosocial stressors at work to exert their effect on cardiometabolic outcomes (37, 38). Inadequate period at risk can also leads to an underestimation of the effect. The present study is consistent with a long-term effect of psychosocial stressors at work on the prevalence of prediabetes, assessed 16 years later, among women.

Results of the present study are in line with previous literature, which reported a stronger and more consistent association between psychosocial stressors at work and T2DM among women (7, 12). A recent systematic review and meta-analysis evaluated the effect of ERI on T2DM incidence. In this review, T2DM incidence was higher in workers exposed to a high ERI (RR=1.24, 95 % CI: 1.08-1.42). Given the small number of individual studies on ERI and T2DM (n=4), results were not stratified by sex. However, this previous meta-analysis also examined the effect of job strain, another work stressor, and reported a stronger and more consistent effect among women. Several hypotheses are put forward to explain the possibility for a sex-specific effect of psychosocial stressors at work on glucose metabolism imbalances. Previous evidence observed that both work-family conflict and total workload (i.e. paid and unpaid work) are higher among women than their male counterparts (39). Moreover, evidence suggests that combining psychosocial stressors at work and high family responsibilities leads to an increase in blood pressure (40). Biological responses to chronic stressors might also differ across sexes. Previous studies showed that women exposed to such stressful situations had steeper psychoneuroendocrine activation (41), including higher cortisol hormone secretion(42) than men. However, further examinations of sex (biological) and gender (socio-cultural) differences are required.

Overall, results did not suggest an association between ERI at work and mean HbA1c concentration. To our knowledge, there was no previous prospective study that has examined the effect of ERI at work on mean HbA1c concentration. However, previous cross-sectional studies found significant associations (33, 35, 43). These conflicting results highlight the need for further quality prospective research on the association between ERI exposure and this important biomarker of glucose metabolism.

Limitations should be acknowledged. First, the present study relied on a convenience sample comprising of workers who were either still active or recently retired at the last follow-up. A comparison analysis showed that the study sample was younger and healthier, on average, than the rest of the cohort (not shown). Therefore, the possibility for a healthy worker effect and effect underestimation cannot be ruled out. Second, HbA1c and prediabetes were assessed at the last follow-up only. Therefore, causal inferences cannot be made. However, previous studies have reported the adverse effect of ERI on T2DM incidence and do not suggest a strong possibility for reverse causation to explain the observed associations. Moreover, the outcome of interest was measured 16 years after assessing ERI exposure, which makes reverse causality unlikely. In the present study, diet was assessed only at the last follow-up, using the alternative healthy eating index (aHEI) among participants who agreed to answer this questionnaire (71.4%). In a sensitivity analysis, we have restricted the study sample to those with available information on diet. Estimates with and without adjustment for diet were identical, minimizing the possibility for residual confounding (not shown). This result should however be interpreted with caution given that the temporal precedence of diet on prediabetes assessment was not respected. Lastly, the cohort involves white-collar workers, which limits generalization of the findings to workers with similar conditions. However, the participants in the present study held a diversity of white-collar occupations such as office workers, technicians, professionals and managers. Moreover, a majority of workers in Organisation for Economic Co-operation and Development (OECD) countries hold white-collar occupations, supporting generalization to a considerable section of the workforce (44).

The present study also had important strengths. First, the association between ERI at work, HbA1c concentration and prediabetes prevalence was examined in a long-term prospective study composed of women and men. Second, a large number of potential confounders were considered, and selected according to a direct acyclic graph (Annex A), in line with recent epidemiological recommendations (45). Third, a sequential adjustment was used to account for the possible role of lifestyle habits as a mediating factor. The estimates did not change after adjusting for lifestyle habits, suggesting that their role as mediators was minor. Lastly, ERI at work, HbA1c and other clinical measurements were performed using validated questionnaires and protocols.

The results of the present study suggest that, among women, ERI at work is associated with a higher prevalence of prediabetes. Prediabetes is on the continuum between normal glucose metabolism and T2DM. Since it is a reversible condition (46), early interventions have the potential to delay and prevent T2DM (47). Screening for psychosocial stressors at work may be relevant to identify patients at risk of developing prediabetes. Physicians and occupational health specialists could help to identify psychosocial stressors at work and cooperate with employers to implement targeted preventive interventions aiming at decreasing these adverse exposures at work. In recent years, non-pharmacological treatment options are considered as the standard of care in prediabetes as well as mild T2DM cases (48). Psychosocial stressors at work are promising targets to intensify non-pharmacological, population-based preventive interventions. Results from the present study suggest that workplace interventions tackling ERI at work may reduce the prevalence of prediabetes among women. Previous studies have shown that psychosocial stressors at work can be reduced through organizational interventions (49) and can provide benefits on workers’ cardiovascular health (50). However, such evidence is scarce and further intervention studies are needed to examine whether reducing psychosocial stressors at work could lead to beneficial effects on prediabetes and T2DM.

## CONCLUSION

This study examined the effect of exposure to effort-reward imbalance on HbA1c concentration and on the prevalence of prediabetes in a prospective cohort. In women, those exposed to ERI at work had an increased prevalence of prediabetes, when compared to unexposed women. Addressing these frequent and modifiable occupational factors could improve the health of workers. Moreover, establishing workplace interventions aiming at decreasing psychosocial stressors at work may be considered as a promising avenue to reduce the prevalence of prediabetes, among women.

## Data Availability

All data produced in the present study are available upon reasonable request to the authors.

## ACKNOWLEDGEMENTS

This research was supported by a grant from the Canadian Institutes of Health Research (grant 57,750). CB was a Canadian Institutes of Health Research Investigator at the time this cohort was initiated, and CR was supported by a Canadian Institutes of Health Research training award when this work was conducted. CR was also supported by training awards from the Fond de Recherche du Quebec en Santé (FRQS) and by VITAM, a research center in sustainable health affiliated with the Centre Intégré Universitaire de Santé et Services Sociaux Capitale-Nationale.

## Annex A

Direct Acyclic Graph (DAG)

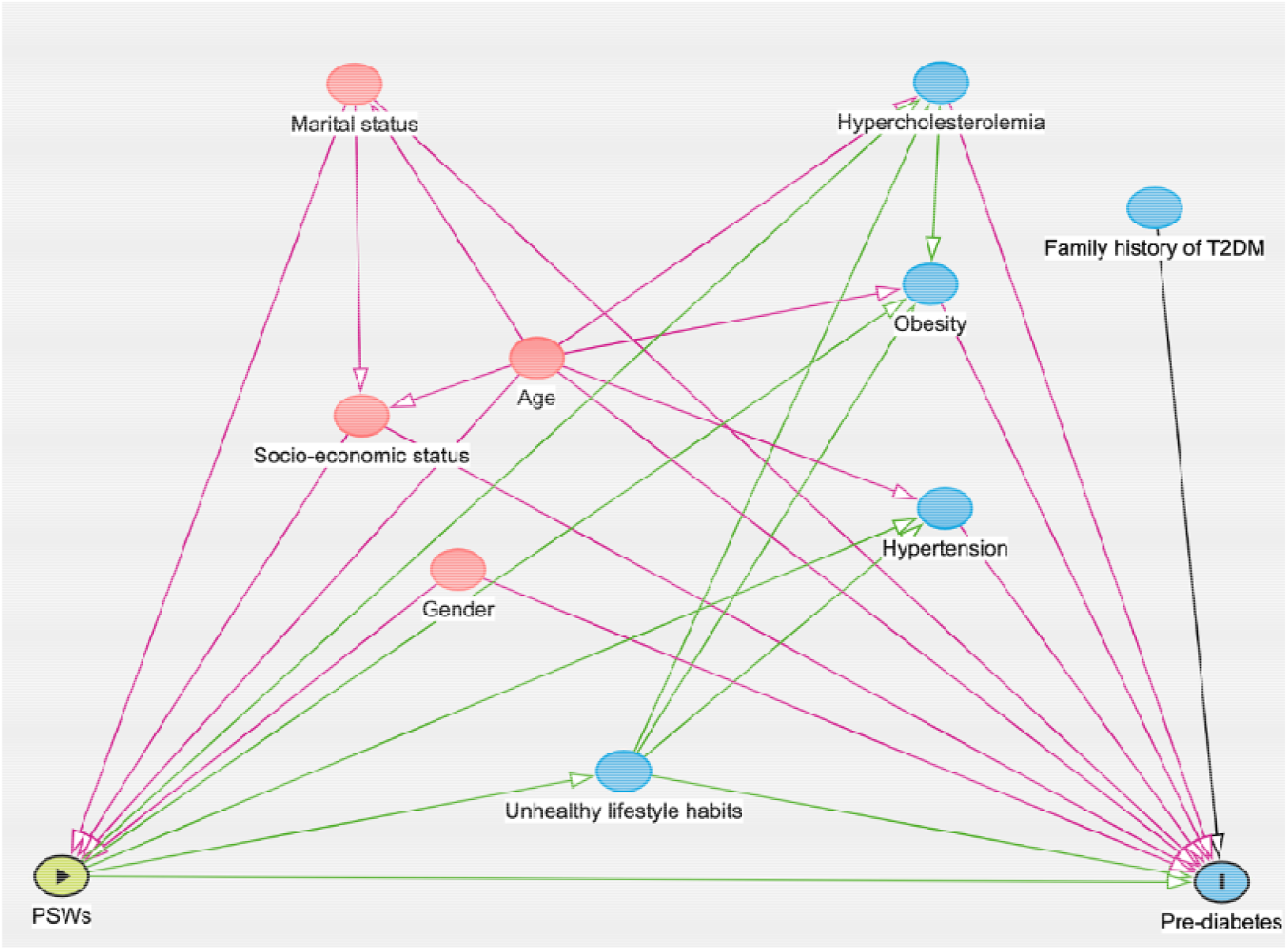

---

**Figure 1.**
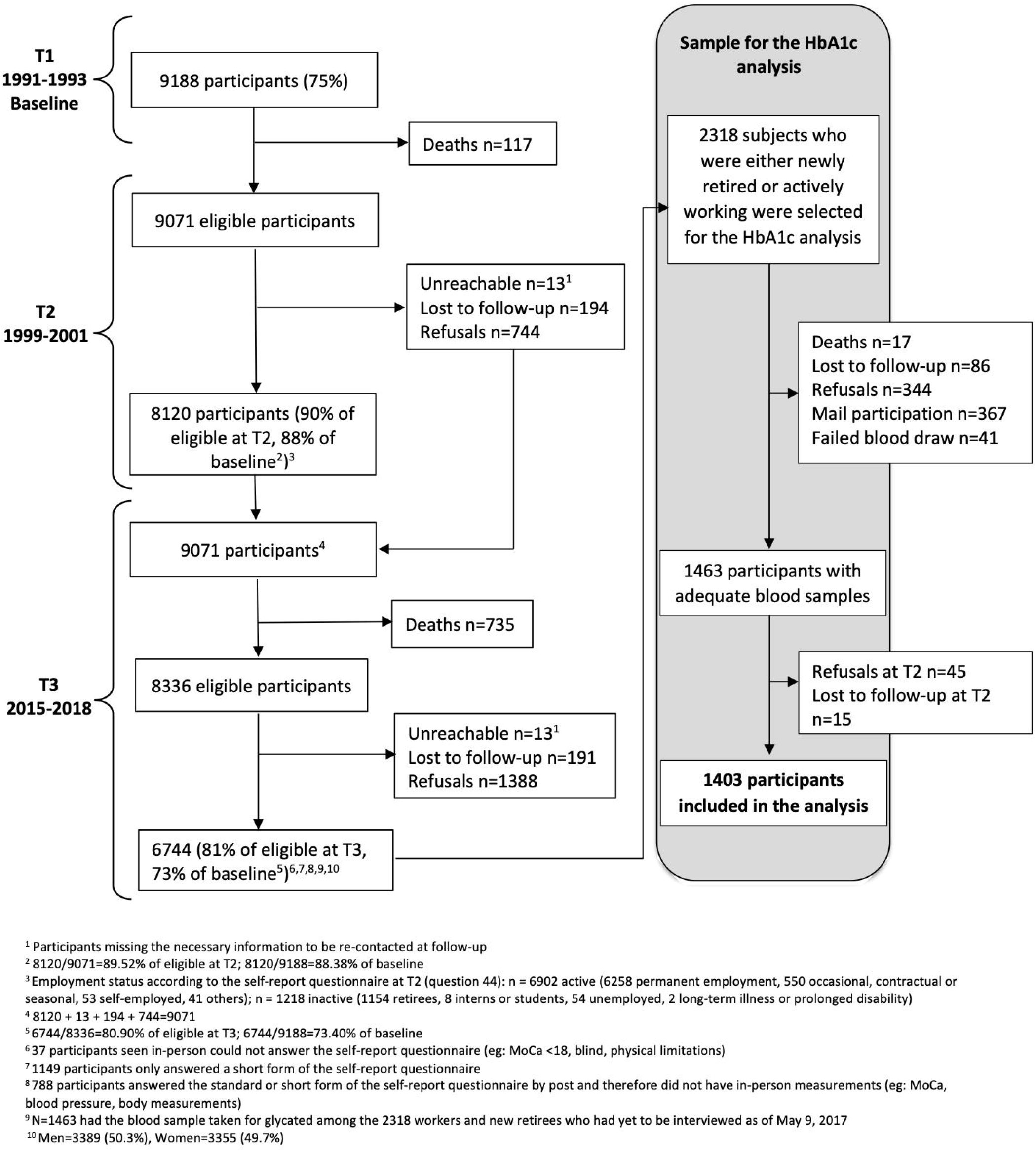
Flowchart of the prospective cohort and the sample for the present study.

